# PREVALENCE OF HEPATITIS B VIRUS SURFACE ANTIGEN AMONG HOSPITAL PATIENTS IN JASHORE CITY, BANGLADESH, 2017-2018

**DOI:** 10.1101/2025.10.01.25337101

**Authors:** Aparajita Kabiraj, Md Deen Islam, Aminur Rahman, Md. Ahsan Habib, Shovon Lal Sarkar, Sabrin Bashar

## Abstract

**Background:** Despite having a huge public health burden, Bangladesh lacks recent national surveillance studies on HBV prevalence and epidemiology. The objective of this study was to understand the epidemiology of HBV infection among the visitors of hospitals in the Jashore region of Bangladesh.

**Methods:** A retrospective type of descriptive study has been conducted among 555 patients from October 2017 to August 2018 in Jashore, Bangladesh.

**Results:** The overall prevalence of hepatitis B surface antigen (HBsAg) seropositivity was 11% in the total sample population. The number of HBV-infected individuals was greater among males compared to females (15% vs 9%, P-value =0.018). The study found that HBV infection was more prevalent among unmarried individuals (15%) compared to those who were single (10%). Participants age ranging from 29-33 years showed highest HBV prevalence (P-value 0.007). After stratifying by sex, only male patients showed significant difference of blood group (P-value 0.019) and age (P-value 0.001) associated with HBV prevalence. Months of sample collection did not show any significant association with HBV prevalence.

**Conclusion:** Our study revealed a higher prevalence of active HBV infection among hospital visitors in Jashore City, especially among males, and those in the younger adult age groups.

## Background

Despite having an effective vaccine and antivirals, over 295 million people around the world are chronically infected with hepatitis B virus (HBV) (1). This chronic hepatitis B (CHB) infection causes liver cirrhosis and hepatocellular carcinoma (HCC) that can lead to death in many cases; approximately 850,000 individuals die annually (2). An astounding portion (about 67.5%) of global HBV-related deaths occur in the WHO South-East Asia region (3, 4).

Geographically located within this region, Bangladesh has a higher prevalence (4-4.5%) of HBV infection in the general population than the global prevalence rate of 3.5% (5). In 2003, the National Expanded Programme on Immunization (EPI) implemented a birth-dose HBV vaccination for the first time in Bangladesh (6,7). In 2019, Bangladesh became one of the first countries in the WHO South-East Asia region to achieve an HBV surface antigen (HBsAg) prevalence of less than 1% among children aged 5 years and below (5, 8).

A notable success was achieved for children compared to adults, mainly due to the inadmissibility of adults to the EPI vaccination. Challenges remain in the lack of sustained funding for HBV immunization programs, limited access to treatment and testing, and the need for nationwide up-to-date data to foster comprehensive national guidelines for viral hepatitis management (9).

The information on the nationwide true prevalence of HBV infection is lacking, mostly due to the lack of routine official reports or surveillance systems. Most of the data found are limited to the government hospitals, while other local private hospitals or diagnostic centers do not share data with the Government (5,10). The last national survey on HBV prevalence was conducted by the Bangladesh Bureau of Statistics in 2014 (4). We need more recent long-term population-based surveillance studies to foster comprehensive national guidelines and policies to execute successful immunization programs.

However, information on the local prevalence of HBV within Bangladesh can better inform the national immunization strategies and overall HBV disease management. To gauge the local HBV disease burden among the general population and to create public health awareness during the blood donation campaign, we conducted a study among the patients who visited two sentinel hospitals in Jashore city, Bangladesh over about one year.The focus of our study is to determine the overall prevalence of hepatitis B surface antigen (HBsAg) in Jashore city.Furthermore, we investigated the influence of age, sex, marital status, blood group, and seasonal variation on the prevalence of HBV infection in the patient population

To address the proposed objectives, we relied on serological testing,one of the standard methods for detecting and monitoring the disease burden of hepatitis B virus (HBV) infection. Serological testing includes mainly the detection of hepatitis B surface antigen (HBsAg), a key serological marker for diagnosis (1), core antigen (HBcAg) and hepatitis B core antibody (anti-HBc). On the other hand, molecular testing includes quantifying HBV DNA viral load directly from patient-derived samples (11).

## Methods

### Study site and population

Two of the privately owned diagnostic hospitals, LABAID Diagnostic Jashore (lat 23.16816218965157, long 89.21034053620886) and Ibn Sina Hospital & Diagnostic Center, Jashore (lat 23.16799204245662, long 89.21049878654559) situated in the center of Jashore City of Bangladesh were selected as they perform diagnosis for HBV. Inclusion criteria were patients only from Lab Aid and Ibn Sina Hospitals of Jashore city with varied sociodemographic factors (eg. Age, sex and marital status) and physiological factors (eg. Blood group). Exclusion criteria were patients living outside of Jashore city.

The scope and purpose of the study were explained to patients who came to the hospital for different treatments. Among those, the interested individuals were invited for blood tests and completing a questionnaire. The data was collected from in-person interviews and by questionnaire supply. A standard questionnaire was used to gather more information about every individual, including data regarding age, gender, group, and marital status. For participants who didn’t know their blood group, a blood grouping test was performed before collecting their samples for the Hepatitis B surface antigen test.

From October 2017 to August 2018, a total of 555 blood samples were randomly collected from visitors to these hospitals. From these individuals written informed consent was sought. The Ethical Review Committee of the Jashore University of Science and Technology (JUST), Bangladesh reviewed and approved this protocol.

### Detection of HBsAg

The laboratory testing was performed in the Dept of Microbiology at JUST, Bangladesh. A solid-phase enzyme-linked immunosorbent assay (ELISA) (cat. EEL140, Thermo Fisher Scientific, USA), based on the “sandwich” principle with the use of microtiter plates, was performed for the detection of HBsAg following published protocols with little modification (12). In summary, 3-4 ml blood samples were collected from individual patients for serum separation. Using a 96-well plate, 50 µl of the negative control was added to well A1, and 50 µl of positive controls to well A2. Serum samples were added to the rest of the wells (A3, A4, A5 etc.) at 50 µl per well followed by 50 µl of enzyme conjugate to each well. After mixing the plate for 30 seconds, the plate was incubated at 37°C for 30 minutes. Followed by 6 repeated washing steps using 350 µl of wash solution, 50 µl of each of substrates A and B were added to each well and incubated at 37°C in the dark for 10 minutes. Then 50 µl of stop solution was added to each well, and mixed gently. Finally, the chemiluminescence was recorded within 20 minutes using a microplate reader at 450 nm or could be read within one day by the naked eye. The colour of the test sample wells was compared to the colour of the positive and negative control wells to determine the HBsAg positivity of a specific sample (patient). The HBsAg One Step Hepatitis B Surface Antigen Test Strip (Serum/Plasma) kit (OnSite HBsAg Rapid Test Strip, Catalog #R0040S, CTK Biotech) was used to further confirm the ELISA-tested positive samples following the kit user manual.

### Statistical Analysis

The data collected was entered in Microsoft Excel and categorized according to qualitative and quantitative variables. The descriptive analysis was analyzed using frequencies, mean, standard deviation, and proportions. Tests of association were performed using chi-square test or Fisher’s exact test based on sample size. The Statistical analysis was performed using STATA v. 13 and all the figures were generated using RStudio v2024.04.1+748. A P-value of <0.05 or <0.1 was considered statistically significant or marginally significant, respectively.

## Results

During the 11 months of surveillance from October 2017 to August 2018,555 patients with varied age groups, blood groups, sex as a biological variable, and marital status were tested for HBsAg (active infection) at two participating hospitals (**Table 1**). Of the patient population, 11% (63/555) showed seropositivity to HBsAg.

**Table 1.** Demographic characteristics of study participants with HBsAg diagnosis results.

| Characteristics | Total<br>N=555 | HBs Ag<br>positive<br>N=63 (11%) | HBs Ag<br>negative<br>N=492 (89%) | P-value<sup>γ</sup> |
| --- | --- | --- | --- | --- |
| **Sex** |  |  |  | 0.018 |
| Male | 223 | 34 (15.25) | 189 (84.75) |  |
| Female | 332 | 29 (8.73) | 303 (91.27) |  |
| **Blood Group** |  |  |  | 0.055 |
| A+ | 97 | 11 (11.34) | 86 (86.66) |  |
| A- | 10 | 1 (10) | 9 (90) |  |
| B+ | 277 | 24 (8.66) | 253 (91.34) |  |
| B- | 8 | 2 (25) | 6 (75) |  |
| AB+ | 34 | 3 (8.82) | 31 (91.18) |  |
| AB- | 6 | 0 (0) | 6 (100) |  |
| O+ | 112 | 18 (16.07) | 94 (83.93) |  |
| O- | 11 | 7 (63.64) | 4 (36.36) |  |
| **Marital Status** |  |  |  | 0.052 |
| Married | 385 | 37 (9.61) | 348 (90.39) |  |
| Unmarried | 170 | 26 (15.29) | 144 (84.71) |  |
| **Months** |  |  |  | 0.451 |
| January | 44 | 2 (4.55) | 42 (95.45) |  |
| February | 52 | 8 (14.55) | 44 (95.45) |  |
| March | 92 | 10 (10.87) | 82 (89.13) |  |
| April | 44 | 5 (11.36) | 39 (88.64) |  |
| May | 19 | 0 (0) | 19 (100) |  |
| June | 44 | 4 (9.09) | 40 (90.91) |  |
| July | 55 | 8 (14.55) | 47 (85.45) |  |
| August | 40 | 3 (7.5) | 37 (92.5) |  |
| September | NA | NA | NA |  |
| October | 53 | 5 (9.43) | 48 (90.57) |  |
| November | 61 | 11 (18.03) | 50 (81.97) |  |
| December | 51 | 7 (13.73) | 44 (86.27) |  |
| **Age groups** |  |  |  | 0.007 |
| <19 | 65 | 7 (10.77) | 58 (89.23) |  |
| 19-23 | 115 | 11 (9.57) | 104 (90.43) |  |
| 24-28 | 134 | 17 (12.69) | 117 (87.31) |  |
| 29-33 | 92 | 24 (26.09) | 68 (73.91) |  |
| 34-40 | 68 | 7 (10.29) | 61 (89.71) |  |
| 41-45 | 24 | 2 (8.33) | 22 (91.67) |  |
| 46-50 | 22 | 3 (13.64) | 19 (86.36) |  |
| >50 | 35 | 1 (2.86) | 34 (97.14) |  |

<sup>γ</sup> P-value generated by chi-square test. Here P-value <0.05 or <0.1 is considered statistically significant or marginally significant, respectively. NA=not available

HBsAg prevalence seemed a year-round phenomenon. HBsAg detection was higher during the winter season (5-18% from November to February) and peaked at November with 18% prevalence. During the pre-monsoon hot season (0-11% from March to May) peaked in April with 11% and during the monsoon/rainy season (8-15% from June to October) peaked in July with 15% prevalence (**Figure 1, Table 1**). However, we did not observe any association between sampling months and sero positivity (P-value= 0.45).

**Figure 1.**
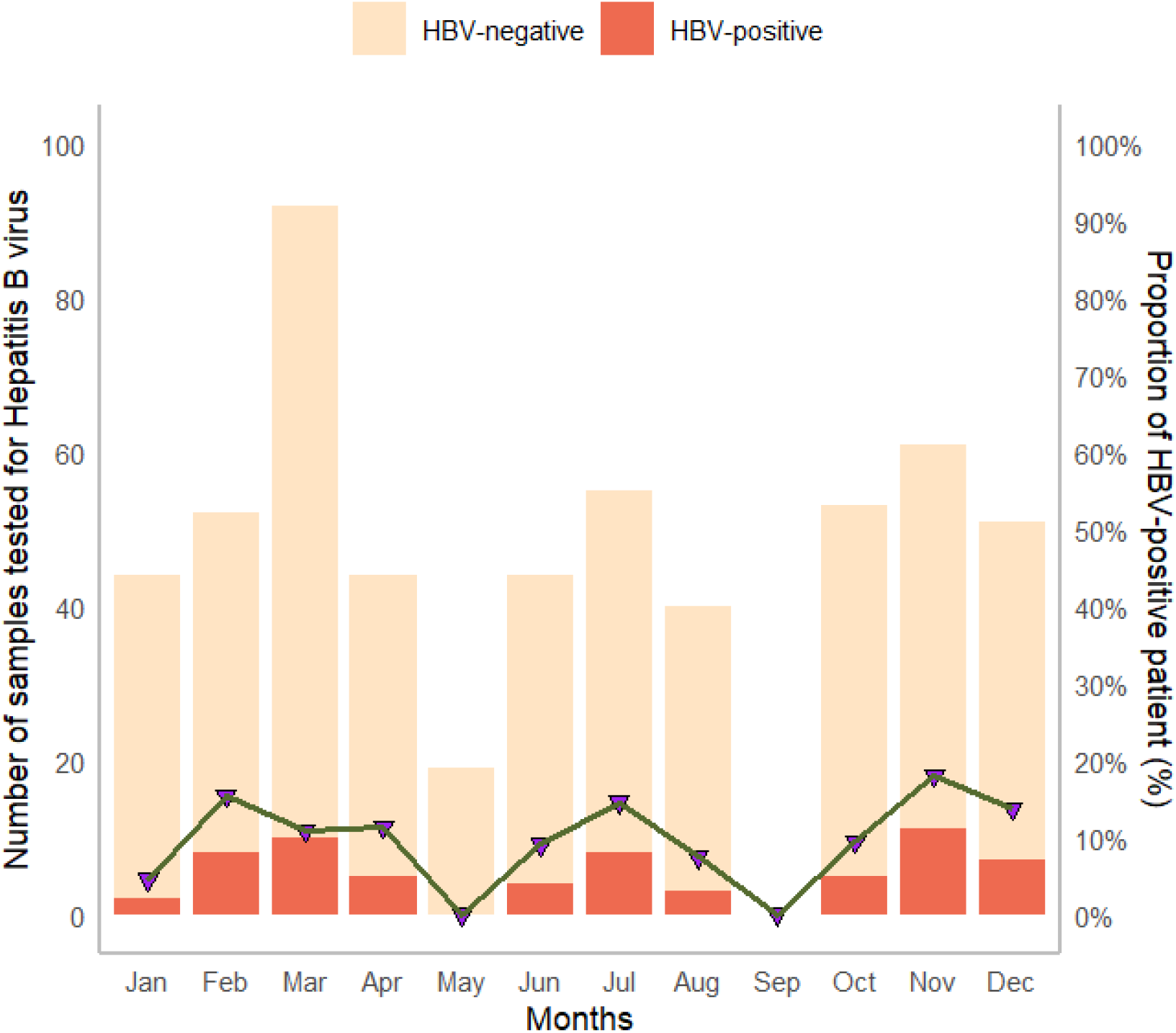
Seasonal fluctuations of HBsAg prevalence among hospital patients in Jashore City, Bangladesh from Oct 2017 to Aug 2018. Along with the y-axis total number of patients tested and the percentage of patients who tested positive for HBsAg each month have been mentioned, respectively. Also, the x-axis represents individual months. We do not have samples information from September 2017

We analyzed whether sex has any effect on the prevalence of HBV infectionis in our study. In our study, the majority of the participants were female (60%, 332/555). However, male participants showed significantly more HBsAg positive than the female patients (male=15.25%, female=8.73%, P-value 0.018, **Table 1**). This result supports the previous works where male showed more HBV than female (5, 13). Therefore we further stratified our data based on sex on each of the biological variables which showed statistically or marginally statiscally significant difference of HBsAg positivity (**Table 2**).

**Table 2.**
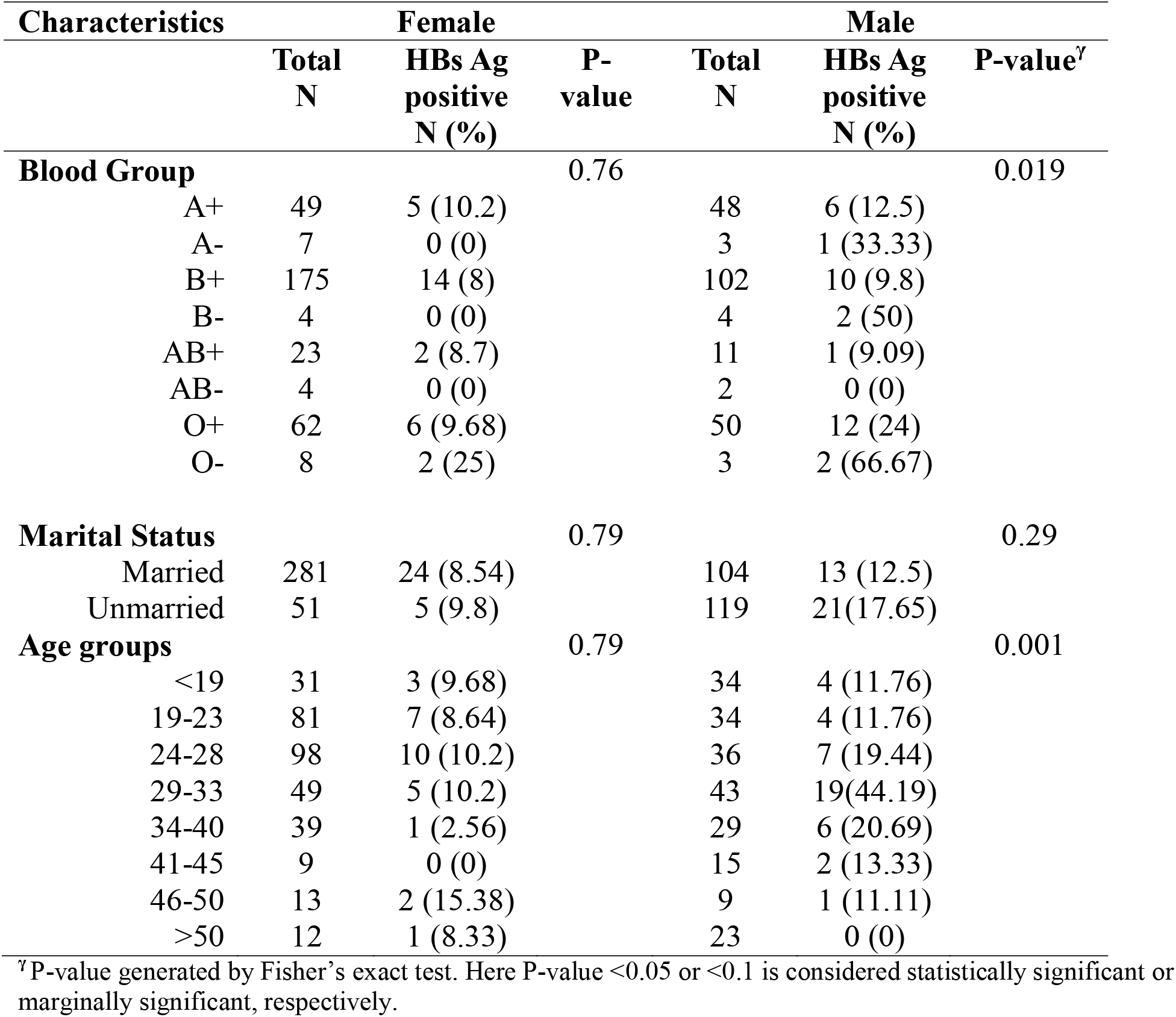
Association of different variables with HBsAg positive stratified by sex.

| Characteristics | Female |  |  | Male |  |  |
| --- | --- | --- | --- | --- | --- | --- |
|  | Total<br>N | HBs Ag<br>positive<br>N (%) | P-value | Total<br>N | HBs Ag<br>positive<br>N (%) | P-value<sup>γ</sup> |
| **Blood Group** |  |  | 0.76 |  |  | 0.019 |
| A+ | 49 | 5 (10.2) |  | 48 | 6 (12.5) |  |
| A- | 7 | 0 (0) |  | 3 | 1 (33.33) |  |
| B+ | 175 | 14 (8) |  | 102 | 10 (9.8) |  |
| B- | 4 | 0 (0) |  | 4 | 2 (50) |  |
| AB+ | 23 | 2 (8.7) |  | 11 | 1 (9.09) |  |
| AB- | 4 | 0 (0) |  | 2 | 0 (0) |  |
| O+ | 62 | 6 (9.68) |  | 50 | 12 (24) |  |
| O- | 8 | 2 (25) |  | 3 | 2 (66.67) |  |
| **Marital Status** |  |  | 0.79 |  |  | 0.29 |
| Married | 281 | 24 (8.54) |  | 104 | 13 (12.5) |  |
| Unmarried | 51 | 5 (9.8) |  | 119 | 21(17.65) |  |
| **Age groups** |  |  | 0.79 |  |  | 0.001 |
| <19 | 31 | 3 (9.68) |  | 34 | 4 (11.76) |  |
| 19-23 | 81 | 7 (8.64) |  | 34 | 4 (11.76) |  |
| 24-28 | 98 | 10 (10.2) |  | 36 | 7 (19.44) |  |
| 29-33 | 49 | 5 (10.2) |  | 43 | 19(44.19) |  |
| 34-40 | 39 | 1 (2.56) |  | 29 | 6 (20.69) |  |
| 41-45 | 9 | 0 (0) |  | 15 | 2 (13.33) |  |
| 46-50 | 13 | 2 (15.38) |  | 9 | 1 (11.11) |  |
| >50 | 12 | 1 (8.33) |  | 23 | 0 (0) |  |

<sup>γ</sup> P-value generated by Fisher's exact test. Here P-value <0.05 or <0.1 is considered statistically significant or marginally significant, respectively.

Among the participants (N=555), the vast majority (61%) were from three age groups ranging from 19 to 33 years old, with the highest prevalence of HBsAg in the age group 29-33 years (26.09%). On the other hand, HBsAg was the least prevalent (3%) in the age group >50 years (2.86%) **(Figure 2(a), Table 1)**. The different age group showed significant difference regarding HBsAg positivity (p-value= 0.007, **Table 1**) After further staritification based on sex, this difference only persist in male patients (P value 0.001, **Table 2**). In male, the age group 29-33 years still showed highest proportion (44.19%) of HBV prevalence than the other age groups.

**Figure 2.**
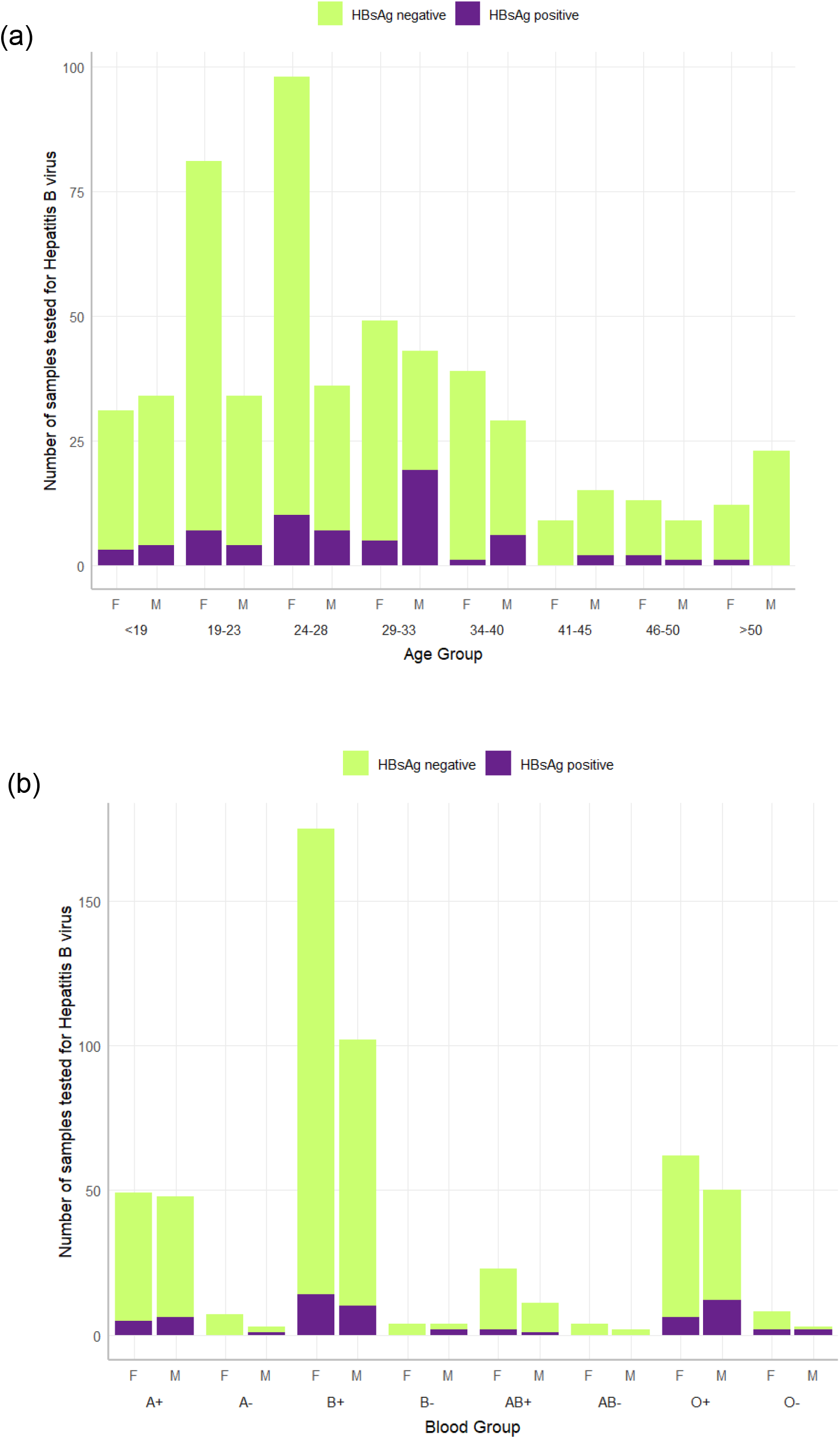
Influence of age and blood groups on HBsAg prevalence between males and females among hospital patients in Jashore City, Bangladesh from Oct 2017 to Aug 2018. Along with the y-axes total number of patients tested and the total HBsAg positive patients. Also, the x-axis represents (a) age groups (years) and (b) ABO blood groups according to sex (male/female (M/F)) with HBsAg prevalence within the groups.

This study confirmed marginally significant association between HBV infection and inherited blood group antigens (p-value= 0.055, **Table 1**). However, the majority of the participants were B+ (50%, 277/555) and the lowest was AB-(1%, 6/555). Among the total participants (N=555), the highest proportion of HBsAg prevalence was found in universal donor group O. For O+ and O-, the proportion were 16.07% and 63.64%, respectively. The lowest prevalence was in the AB-group (0%, 0/6) (**Figure 2, Table 1**). In male patients only, blood group antigen showed significant association with HBV infection (P-value 0.019, **Table 2**). We did not observe any significance difference of blood group and prevalence of HBV infection in female participants.

Among the respondents (N=555), about 69% (385) were married and 31% (170) were unmarried (**Figure 3)**. Marital status showed marginally significant association with HBV infection (P-value 0.05) (**Table 1**). Among the patients, the higher prevalence of HBsAg was found in the unmarried group (15.29%) compared to the married group (9.61%) (P value=0.05). This factor did not show any significant association with HBV once we staritified our data based on sex (P value >0.1, **Table 2**).

**Figure 3.**
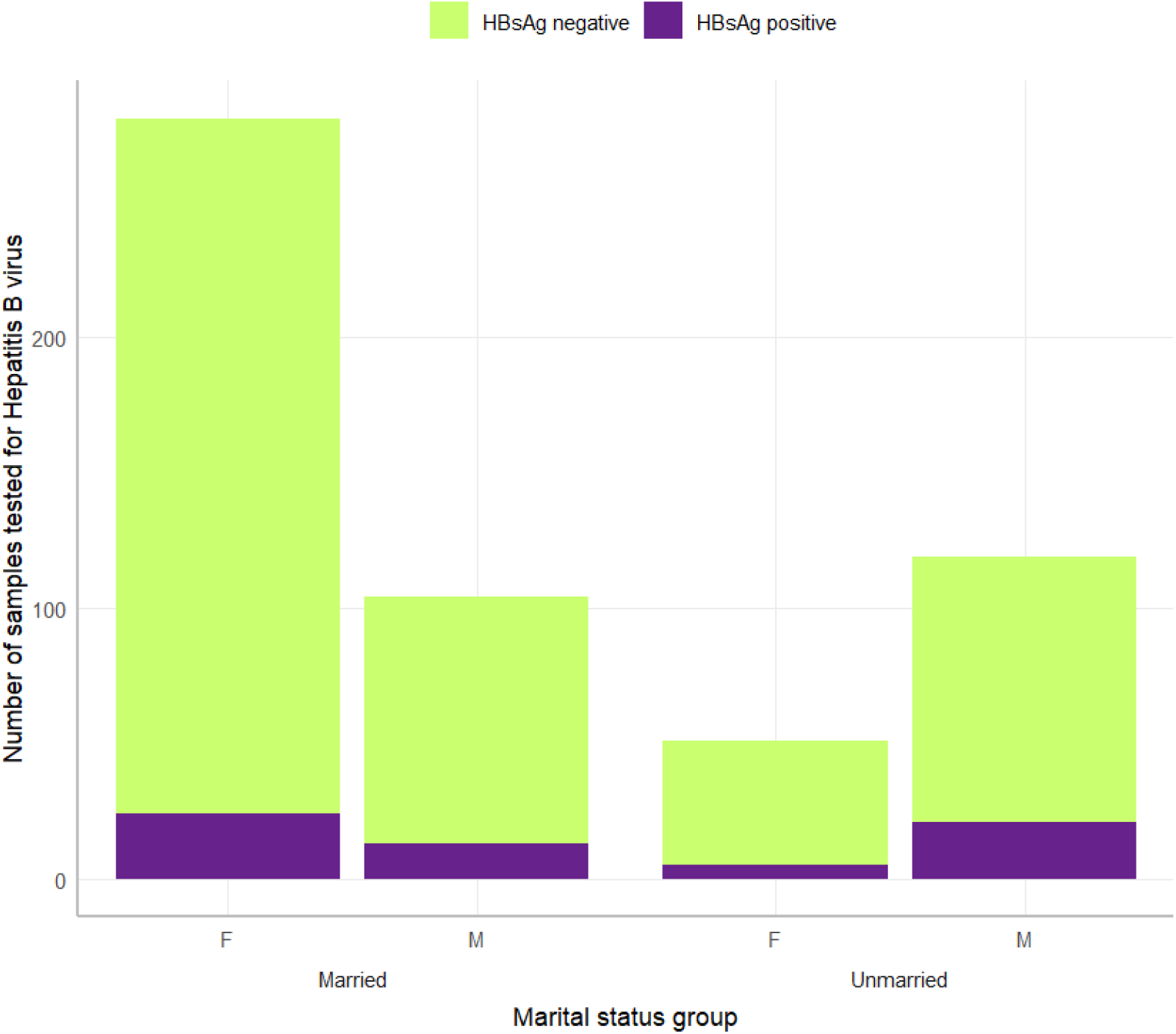
Influence of marital status on HBsAg prevalence between males and females among hospital patients in Jashore City, Bangladesh from Oct 2017 to Aug 2018. Along the y-axes total number of patients tested and the total HBsAg positive patients. Also, the x-axis represents marital status groups (married vs unmarried) and the distribution of male/female (M/F) with HBsAg prevalence within the groups.

## Discussion

According to our study, 11% of the total population is positive for Hepatitis B virus surface antigen, which indicates a high prevalence rate (i.e. ≥8% prevalence) (14). A study showed pooled estimated prevalence of HBV infection in Bangladeshi population from 1995 to 2017 was 4.0% (15). Studies conducted in patients of Dhaka (Bangladesh) observed that the prevalence of Hepatitis B virus with very high percentage that ranges from 8.6 to 27.2% (16, 17). These ranges were so high probably these studies were performed before the expanded program of vaccination (EPI) by the Bangladesh government since 2003 (18). Another study reported that the prevalence of the hepatitis B virus among the general population of Bangladesh is high (8.1 %) (19). Laskar et al. (20), recorded Hepatitis B virus cases at a very lower percentage ( 0.8% positive) by studying among the healthy female students of Viqarunnessa Noon Girls’ School (VNGS), Dhaka. This study reports that lower percentage of Hepatitis B virus cases. The reason for low prevalence could be all participants were healthy with high scocioeconomic status. Moreover, Islam et al. (13) reported an intermediate endemicity of Hepatitis B virus cases by studying among university students in Bangladesh (5% seropositivity). However, these studies are mostly performed in Dhaka City, and unable to define the prevalence of Hepatitis B virus cases in Jashore city.

The prevalence of Hepatitis B virus varies from region to region and different population subgroups. This inconsistency may be due to the extent of awareness, socioeconomic classes, and level of effort to avoid infection by the mass population (5). Our study conducted in Lab Aid and Ibn Sina Hospital which are situated in the center of Jashore city. Most of the people who visit here for treatment are from nearby rural areas and live under low socioeconomic status. Socioeconomic factor was found to be a crucial factor for the prevalence of Hepatitis B (21). Moreover, we collected the blood samples from hospital patients. Therefore, they are immunologically compromised group.

In our study, higher proportion of HBsAg positive was detected in patients from age 29-33 years (24 out of 92, 26%), 24-28 years (17 out of 134, 13%) and 19-23 years (11 out of 115, 10%) age groups than the much younger or elder aged groups. This is contrarory to the previous studies in Bangladesh which showed prevalence of hepatitis B was higher in age <25 years than >25 years or much younger age (5, 10). In our study, the young adult aged groups were born before the national introduction of the hepatitis B vaccine program in Bangladesh (began around 2003). As a result, this cohort may have had incomplete vaccination coverage during infancy, leaving them at higher risk compared to younger cohorts who might have benefitted from routine immunization. Moreover, individuals in these age ranges are generally more socially active, with higher likelihood of exposure to risk behaviors such as unsafe sexual practices, use of intravenous drugs, or unsafe medical interventions, which have been found to contribute significantly to HBV transmission. It is noteworthy to mention that the drug addiction rate and promiscuous sexual activity are high among young people compared to elderly people in Bangladesh (22).

Moreover, we found unmarried people are more prone to infection with the hepatitis B virus than married people which is in contrary to the previous studies conducted in Bangladesh (5,13). Improper sexual practice between spouses contributes to the spread of HBV through semen and vaginal secretion (5). In contrary, Ghadir et al. found singles and divorced individuals had higher HBV rates in Iran, suggesting variability in sexual networking and exposure risks (23). Moreover, Knowledge about HBV transmission can differ by marital status; unmarried adults may have less access to family-based health education or screening. After stratifying by sex, marital status did not show a significant association with HBV infection in the current dataset, which suggests possible effect modification.

The prevalence of HBsAg has been found higher among males compared to females globally but it varies county to country or associated with other lifestyle varibles (24). Most of the studies conducted on the prevalence of HBV infection in Bangladesh, which provides gender-related data (now biologically defined as sex), are in the same line with this fact. The reason behind the high rate of infection among male in Bangladesh was found associated occupational role (working outside of home), less knowledge of HBV infection, and blood transfusion (25). Studies also showed that males with lower socioeconomic status in Bangladesh underwent circumcision with unregistered healthcare providers which might be associated with transmission of this pathogen (26, 27).

We found marginally significant association among ABO blood groups and the presence of HBsAg and significance only persists in male patients after stratification. Recent literature presents mixed findings on the association between ABO blood groups and hepatitis B surface antigen (HBsAg) positivity, with evidence of regional and sex-specific significance. A systematic review and meta-analysis involving 241,868 HBV-infected individuals reported that blood group B is associated with a lower risk of HBV infection, whereas blood group O may confer a slightly increased risk, particularly in high-endemic areas and Asian populations (28). Notably, the association was more pronounced and statistically significant in male patients after stratification, as seen in several large cohort studies, including research on Chinese chronic hepatitis B (CHB) patients, where non-O blood groups were linked to increased disease severity and risk of progression, especially among men (29).

Additionally, research from Nigeria notes a higher HBV risk in individuals with B antigen lacking Rh D antigen, though ABO associations alone were often marginally significant and varied across populations (30). We also observed more HBV prevalence in Rh-patients with blood group B (2 HBsAg positive out of 8 patients, 25%) and O (7 HBsAg postive out of 11 patients, 63.64%). While some investigations fail to find a consistent link, a growing body of meta-analytic and population-based evidence supports the notion that ABO blood group influences HBsAg prevalence, with significance commonly persisting in male subgroups (31, 32). Like all other studies, our study also has some limitations. The information regarding participants was only limited to gender, age and marital status, but did not include any history of infection, blood transfusion, drug addiction, previous vaccination, socioeconomic status, etc. However, this pilot study gives an idea of the prevalence of Hepatitis B in Jashore City which will be applicable towards public health concern.

## Conclusion

In conclusion, the overall prevalence of HBV infection among the general population of Jashore City in Bangladesh is quite high (about 11%). Our findings provide important baseline data on the overall prevalence of HBsAg within the Jashore area that might inform better strategies for vaccination in adults and blood transfusion. Additional studies, including country-wide, community-based and hospital-based surveillance are necessary to understand the comprehensive public health burden of chronic HBV in Bangladesh.

## Data Availability

All data produced in the present study are available upon reasonable request to the authors

## Acknowledgment

We appreciate the study participants for their time and support. We are also grateful for the support and resources provided by our surveillance hospital sites and the Dept of Microbiology at the Jashore University of Science and Technology (JUST).

